# COVID-19 serology at population scale: SARS-CoV-2-specific antibody responses in saliva

**DOI:** 10.1101/2020.05.24.20112300

**Authors:** Pranay R. Randad, Nora Pisanic, Kate Kruczynski, Yukari C. Manabe, David Thomas, Andrew Pekosz, Sabra L. Klein, Michael J. Betenbaugh, William A. Clarke, Oliver Laeyendecker, Patrizio P. Caturegli, H. Benjamin Larman, Barbara Detrick, Jessica K. Fairley, Amy C. Sherman, Nadine Rouphael, Srilatha Edupuganti, Douglas A. Granger, Steve W. Granger, Matthew Collins, Christopher D. Heaney

**Author notes:** Contributed equally (Pranay R. Randad, Nora Pisanic). **Author Contributions**: All authors reviewed and edited all sections of the article. P.R.R. and N.P. wrote the first draft of the manuscript. K.K. and N.P handled laboratory logistics and generated data. N.P. and P.R.R. analyzed and summarized the data. A.P. provided input on study design and edited the manuscript. M.J.B., S.W.G., D.A.G. provided input on antigen selection, assay design, and interpretation of results. Y.C.M and D.T. provided input on study design and interpretation of results. B.D. provided input on interpretation of results. W.A.C, O.L., P.P.C., and B.L. shared samples and data for the analysis and provided input on interpretation of results. M.H.C developed project concept. M.H.C, N.R., J.F., and A.C.S. led and coordinated specimen collection efforts and reviewed and edited the article. C.D.H. developed project concept and guided the laboratory work. **Sources of funding**: Funding for this study was provided by the Johns Hopkins University Provost’s Office and the FIA Foundation. P.R.R., N.P., K.K., and C.D.H. were supported by a gift from the GRACE Communications Foundation. C.D.H., N.P., and B.D. were additionally supported by National Institute of Allergy and Infectious Diseases (NIAID) grants R21AI139784 and R43AI141265, and National Institute of Environmental Health Sciences (NIEHS) grant R01ES026973. C.D.H. was also supported by NIAID grant R01AI130066 and NIH grant U24OD023382. A.P. and S.L.K. were supported by NIH/NIAID Center of Excellence in Influenza Research and Surveillance contract HHS N2772201400007C. O.L. was supported by the Division of Intramural Research, NIAID. The funders had no role in study design, data analysis, decision to publish, or preparation of the manuscript. **Ethical Statement**: This study has been approved by the Johns Hopkins Bloomberg School of Public Health Institutional Review Board (IRB) (IRB No. IRB00012253) Johns Hopkins Medicine IRB (IRB No. IRB00247886) and by the Emory University Institutional Review Board (IRB No. 00110683).

## Abstract

Non-invasive SARS-CoV-2 antibody testing is urgently needed to estimate the incidence and prevalence of SARS-CoV-2 infection at the general population level. Precise knowledge of population immunity could allow government bodies to make informed decisions about how and when to relax stay-at-home directives and to reopen the economy. We hypothesized that salivary antibodies to SARS-CoV-2 could serve as a non-invasive alternative to serological testing for widespread monitoring of SARS-CoV-2 infection throughout the population. We developed a multiplex SARS-CoV-2 antibody immunoassay based on Luminex technology and tested 167 saliva and 324 serum samples, including 134 and 118 negative saliva and serum samples, respectively, collected before the COVID-19 pandemic, and 33 saliva and 206 serum samples from participants with RT-PCR-confirmed SARS-CoV-2 infection. We evaluated the correlation of results obtained in saliva vs. serum and determined the sensitivity and specificity for each diagnostic media, stratified by antibody isotype, for detection of SARS-CoV-2 infection based on COVID-19 case designation for all specimens. Matched serum and saliva SARS-CoV-2 antigen-specific IgG responses were significantly correlated. Within the 10-plex SARS-CoV-2 panel, the salivary anti-nucleocapsid (N) protein IgG response resulted in the highest sensitivity for detecting prior SARS-CoV-2 infection (100% sensitivity at ≥10 days post-SARS-CoV-2 symptom onset). The salivary anti-receptor binding domain (RBD) IgG response resulted in 100% specificity. Among individuals with SARS-CoV-2 infection confirmed with RT-PCR, the temporal kinetics of IgG, IgA, and IgM in saliva were consistent with those observed in serum. SARS-CoV-2 appears to trigger a humoral immune response resulting in the almost simultaneous rise of IgG, IgM and IgA levels both in serum and in saliva, mirroring responses consistent with the stimulation of existing, cross-reactive B cells. SARS-CoV-2 antibody testing in saliva can play a critically important role in large-scale “sero”-surveillance to address key public health priorities and guide policy and decision-making for COVID-19.

**40-word summary:** A multiplex immunoassay to detect SARS-CoV-2-specific antibodies in saliva performs with high diagnostic accuracy as early as ten days post-COVID-19 symptom onset. Highly sensitive and specific salivary COVID-19 antibody assays could advance broad immuno-surveillance goals in the USA and globally.

## Introduction

The coronavirus disease 2019 (COVID-19) pandemic, caused by severe acute respiratory syndrome virus 2 (SARS-CoV-2), has caused >5.4 million COVID-19 cases and >344,000 deaths, as of May 24, 2020, involving all populated continents.^1^ The USA accounts for >1.6 million COVID-19 cases and >97,000 deaths and the outbreak has expanded from urban to rural areas of the country.^1^ There is a critical need to perform broad-scale population-based testing to improve COVID-19 prevention and control efforts. Some have even recommended national testing at repeated time points to improve understanding of the spatio-temporal dynamics of transmission, infection, and herd immunity.^2,3^ Currently, population-level antibody testing is largely performed using blood, with preliminary seroprevalence study estimates ranging from 2.8.% in Santa Clara County, California,^4^ 4.65% in Los Angeles County, California,^5^ 21% in New York City,^6^ 11.5% in Robbio Italy,^7^ and 14% in Gangelt, Germany.^8^ Achieving such comprehensive national testing goals will be challenging by relying only on traditional blood-based diagnostic specimens as these may be considered too invasive, uncomfortable, or unacceptable, particularly among vulnerable and susceptible groups.^9-12^

In addition to molecular COVID-19 diagnostics, accurate serological tests can identify individuals who have mounted an antibody response to SARS-CoV-2 infection. These tests are needed in platforms that can be deployed in large numbers to describe changes in population level immunity at different geographical scales and over time. Such serological testing could guide “back-to-work” risk mitigation strategies^2,3^, particularly if evidence continues to emerge suggesting that robust SARS-CoV-2 antibody responses might confer protection from repeated infection.^13,14^

Saliva harvested from the space between the gums and the teeth is enriched with gingival crevicular fluid (GCF). The composition of GCF (hereafter referred to as “saliva”) resembles that of serum, and is enriched with antibodies.^15-24^ Thus, sampling saliva with an appropriate collection method is an attractive non-invasive approach for antibody-based diagnostic techniques. We have previously demonstrated the utility of saliva-based serology testing for the diagnosis, surveillance, and study of infection by multiple viral pathogens.^21,22^ Development of improved antibody assays to detect prior infection with SARS-CoV-2 has been identified as one of the top unmet needs in the ongoing COVID-19 pandemic response.^2,3^ Precise knowledge of SARS-CoV-2 infection at the individual level can potentially inform clinical decision-making, whereas at the population level, precise knowledge of prior infection, immunity, and attack rates (particularly asymptomatic infection) is needed to prioritize risk management decision-making about social distancing, treatments, and vaccination (once the latter two become available).^25^ If saliva can support measurements of both the presence of SARS-CoV-2 RNA^26-28^ as well as antibodies against SARS-CoV-2, this sample type could provide an important opportunity to monitor individual and population-level SARS-CoV-2 transmission, infection, and immunity dynamics over place and time.

Prior studies have shown that antibodies to SARS-CoV-2 nucleocapsid protein (N), spike protein (S), and the receptor binding domain (RBD) are elevated in serum around 10-18 days following SARS-CoV-2 infection.^14,29-32^ Many ELISA, point-of-care (POC), and lateral flow IgG assays for detecting prior SARS-CoV-2 infection that are currently available show a wide range in diagnostic performance. The sensitivity of the assays improves when samples are collected later after the onset of infection, from <20% sensitivity at <5 days to approximately 100% sensitivity at 17 to 20 days from symptom onset.^33-35^

In this study, we aimed to determine whether salivary SARS-CoV-2-specific antibody responses would identify prior SARS-CoV-2 infection with similar sensitivity and specificity as serum and whether salivary antibody testing would reflect the temporal profiles observed in serum. The objectives of this study were: (1) to develop and validate a multiplex bead-based immunoassay for detection of SARS-CoV-2-specific IgG, IgA, and IgM responses; (2) to describe the assay performance using saliva compared to using serum specimens; (3) to identify SARS-CoV-2 antigens that could result in high sensitivity and specificity to identify antibody responses to prior SARS-CoV-2 infection; and (4) to compare the antibody kinetics in saliva to those in serum by time since onset of COVID-19 symptoms.

## Methods

### Sources of saliva and serum

Saliva and serum samples were provided by collaborators from Emory University from patients in three settings: 1) PCR-confirmed COVID-19 cases while admitted to the hospital; 2) confirmed COVID-19 cases we invited to donate specimens after recovering from their acute illness; and 3) patients with symptoms consistent with COVID-19 being tested at an ambulatory testing center donated specimens at the time of testing and/or at a follow-up convalescent phase research visit. Collaborators at Johns Hopkins University provided: 1) serum samples from patients presenting with COVID-19-like symptoms such as fever, cough, dyspnea who were recruited in both inpatient and outpatient clinical cares sites; and 2) negative saliva and serum samples collected prior to the COVID-19 pandemic. Participants provided verbal and / or written informed consent and provided saliva and blood specimens for analysis. Whenever possible, remnant clinical blood specimens were used. Basic data on days since symptom onset were recorded for all participants as were results of COVID-19 molecular testing. Participation in these studies was voluntary and the study protocols have been approved by the respective Institutional Review Boards.

### Saliva and blood sample collection

Saliva samples were collected by instructing participants to gently brush their gum line with an Oracol S14 saliva collection device (Malvern Medical Developments, UK) for 1-2 minutes, or until saturation. This saliva collection method specifically harvests GCF, which is enriched with primarily IgG antibody derived from serum.^18^ The saturated sponge was then inserted into the storage tube, capped, and stored at 4°C until processing whenever possible. Saliva was separated from the Oracol S14 swabs through centrifugation (10 min at 1,500 g) and transferred into the attached 2 mL cryovial. Samples were heat-inactivated at 60°C for 30 minutes and then shipped to the lab on dry ice. Blood samples were collected into ACD (acid, citrate, dextrose) or serum separator tubes (SST) and processed according to each clinical lab’s procedure. Plasma/serum was also heat inactivated at 60°C for 30 minutes, aliquoted into 2mL cryovials, and stored at ≤20°C until analyzed. Only de-identified serum or plasma and saliva aliquots including limited metadata (days since symptom onset and SARS-CoV-2 RT-PCR status [ever positive or negative]) were shared for this study.

### Multiplex magnetic microparticle (“bead”)-based SARS-CoV-2 saliva immunoassay

Ten SARS-CoV-2 antigens were obtained commercially or from collaborators at Icahn School of Medicine at Mount Sinai (Table 1).^36^ This included four SARS-CoV-2 receptor binding domain (RBD), one ectodomain (ECD) protein containing the S1 and S2 subunit of the spike protein, two S1 subunits, one S2 subunit, and two N proteins. Each SARS-CoV-2 antigen, along with one SARS-CoV-1 antigen (NAC SARS 2002 N) and one human coronavirus (hCoV)-229E antigen (Sino Biol. hCoV 229E ECD), were covalently coupled to magnetic microparticles (MagPlex microspheres, Luminex) as described previously (**Table 1**).^21,22^ Along with a control bead, conjugated with bovine serum albumin (BSA), the multiplex panel included a total of 13 bead sets (10 bead sets coupled to SARS-CoV-2 antigens, one to SARS-CoV-1 antigen, one to hCoV-229E antigen, and one control bead coupled to BSA). Coupling of antigens to beads was confirmed using antibody against the antigen or against the tag (e.g. anti-His(6) tag antibody), if present (**Table 1**), followed by a species-specific R-phycoerythrin (PE)-labelled antibody and was considered successful if the median fluorescence intensity (MFI [a.u.]) was >10,000 at 1 μg/mL of antigen-specific antibody (except the BSA-conjugated bead set). Saliva samples were centrifuged (5 minutes at 20,000*g*, 20°C), and 10 μL of saliva supernatant was added to 40 μL of assay buffer (phosphate-buffered saline with 0.05% Tween20, 0.02% sodium azide and 1% BSA) containing 1,500 beads of each bead set per microplate well. The plate was covered and incubated at room temperature for 1 hour on a plate shaker at 500 rpm. Beads were washed twice with 200 μ PBST and 50 μ of PE-labeled anti-human IgG, IgA or IgM diluted 1:100 in assay buffer were added, and the plate was incubated again for 1 hour on a plate shaker at 500 rpm. Beads were washed as above and then suspended in 100 μL of assay buffer. Finally, the MFI of each bead set was measured on a Bio-Plex® immunoassay instrument (Bio-Rad Laboratories, Hercules, CA). The same protocol was used for serum and plasma samples, except that serum and plasma samples were tested at a final dilution of 1:1000 in bead mix and assay buffer compared to a final dilution of 1:5 for saliva. A subset of 47 saliva samples were tested in duplicate and in a masked fashion to determine intra-assay variability (same 96 well plate) and inter-assay variability (different 96 well plates on different days), and at least 2 blanks (assay buffer) were included on each plate for background fluorescence subtraction.

**Table 1.** Antigens and antibodies used to develop the multiplex bead-based assay.

| <u>Source*</u> | <u>Pathogen</u> | <u>Antigen<sup>#</sup></u> | <u>Tag</u> | <u>Antibody<sup>^</sup></u> | <u>Abbreviation</u> | <u>Antigen Catalog No.</u> | <u>Antibody Catalog No.</u> |
| --- | --- | --- | --- | --- | --- | --- | --- |
| GenScript |  | N | his | anti-Gen N | GenScript N | Z03480 | A02039 |
| Sino Biol. |  | N | his | anti-Sino N | Sino Biol. N | 40588-V08B | A02039 |
| Sino Biol. |  | ECD (S1+S2) | his | anti-Sino RBD | Sino Biol. ECD (S1+S2) | 40589-V08B1 | A02038 |
| Sino Biol. |  | RBD | his | anti-Sino RBD | Sino Biol. RBD | 40592-V08H | 40592-T62 |
| Mt. Sinai |  | RBD | his | anti-Sino RBD | Mt. Sinai RBD | *Amanat F., et al* | 40592-T62 |
| GenScript | SARS-CoV-2 | RBD | his | anti-Sino RBD | Sino Biol. RBD (h) | Z03479 | 40592-T62 |
| GenScript |  | RBD | his | anti-Sino RBD | Sino Biol. RBD (i) | Z03483 | 40592-T62 |
| GenScript |  | S1 | N/A | anti-Gen S | GenScript S1 | Z03501 | A02038 |
| NAC |  | S1 | shFc | anti-Sheep Fc | NAC S1 | REC31806 | 313-005-046 |
| NAC |  | S2 | shFc | anti-Sheep Fc | NAC S2 | REC31807 | 313-005-046 |
| NAC | SARS-CoV-1 | SARS CoV N | his | anti-his | NAC SARS 2002 N | REC31744 | MA121315 |
| Sino Biol. | hCoV-229E | 229E ECD | his | anti-his | Sino Biol. hCoV 229E ECD | 40605-V08B | MA121315 |

\*Sino Biol.: Sino Biological; NAC: Native Antigen Company
<sup>#</sup>N: nucleocapsid protein; ECD: ectodomain (S1 + S2 subunit of spike protein); RBD: receptor binding domain
<sup>^</sup>Corresponding IgG antibody used for confirmation

### Statistical analysis

The median fluorescence intensity (MFI) measured using the BSA beads was subtracted from each blank-subtracted antigen-specific MFI signal for each sample to account for non-specific binding of antibodies to beads. The average MFI was used for samples that were tested in duplicate (n=47) or triplicate. Wilcoxon-Mann-Whitney test was used to compare the median MFI between samples collected <10 days post symptom onset and negatives, and between samples collected ≥10 days post symptom onset and negatives, for each antigen in the multiplex. The average intra- and inter-assay variability was evaluated by determining the coefficient of variation (CV%) of a subset of 47 samples that were tested in duplicate (intra) and on different days and plates (inter). Pearson’s correlation was used to determine the correlation between antigen-specific IgG, IgA, and IgM MFI in matched saliva and serum / plasma samples collected from the same person at the same time point (n=28). The average MFI of all saliva samples from known uninfected individuals (pre-Covid-19) plus three standard deviations for each antigen-specific IgG, IgA, and IgM were used to establish the cut-off values for a negative result. The corresponding procedure was used for serum samples. Because the prior hCoV infection status for saliva and serum samples was not known, the MFI cut-off values were not calculated for anti-Sino Biol. hCoV 229E ECD IgG, IgA, and IgM. Sensitivity and specificity for detecting samples from confirmed RT-PCR positive individuals and for samples from individuals obtained prior to the COVID-19 pandemic were determined for each antigen/isotype pair (IgG, IgM and IgA) in saliva and in serum. Locally weighted regression (LOESS) was used to visualize and compare the temporal kinetics of saliva and serum antigen-specific IgG, IgA, and IgM responses among individuals with RT-PCR confirmed prior SARS-CoV-2 infection post symptom onset.

## Results

### Saliva and serum samples

A total of 33 saliva samples and 206 serum samples were collected from 33 and 59 individuals, respectively, with RT-PCR confirmed prior SARS-CoV-2 infection (Table 2). Information on days post symptom onset was collected for each positive participant. A total of 134 saliva samples (from 2012 to early 2019) and 112 serum samples (from 2016)^37^ were collected from participants enrolled in cohort studies prior to the start of the COVID-19 pandemic and were designated as negative samples (pre-COVID-19 pandemic) (Table 2).

**Table 2.** Saliva and serum samples.

|  | <u>Saliva</u> |  | <u>Serum</u> |  |
| --- | --- | --- | --- | --- |
|  | Participants | Samples | Participants | Samples |
|  | n (%) | n (%) | n (%) | n (%) |
| **All samples** | **150 (100)** | **167 (100)** | **171 (100)** | **324 (100)** |
| SARS-CoV-2 PCR positive | 33 (22.0) | 33 (19.8) | 59 (34.5) | 206 (63.6) |
| SARS-CoV-2 PCR negative | 117 (78.0) | 134 (80.2) | 112 (65.5) | 118 (36.4) |
| ***Matched saliva-serum samples*** | **28 (100)** | **28 (100)** | - | - |
| SARS-CoV-2 PCR positive | 22 (78.6) | 22 (78.6) | - | - |
| SARS-CoV-2 PCR negative | 6 (21.4) | 6 (21.4) | - | - |

### SARS-CoV-2 antigen-specific IgG, IgA, and IgM cut-off values

The multiplex immunoassay, comprised of ten SARS-CoV-2 antigens (2 N proteins, 1 ECD protein, four RBD proteins, two S1 subunits, and one S2 subunit), one SARS-CoV-1 antigen (NAC SARS CoV 2002 N), and one hCoV-229E antigen (Sino Biol. hCoV 229E ECD) was used to test a total of 167 saliva samples from 150 individuals and 324 serum samples from 171 individuals. The range, median, mean, standard deviation, and derived MFI cut off value for each saliva and serum SARS-CoV-2 antigen-specific IgG, IgA, and IgM stratified by negative samples, samples collected <10 days, and ≥10 days post SARS-CoV-2 symptom onset are provided in **Supplementary Table 1** and **Supplementary Table 2**. Saliva collected at ≥10 days post symptom onset had significantly elevated IgG levels (median MFI) against all SARS-CoV-2 antigens compared to negative saliva samples (**Supplementary Table 1**). Serum collected at ≥10 days post symptom onset had significantly elevated IgG, IgA, and IgM levels (median MFI) against all SARS-CoV-2 antigens compared to negative sera.

### Correlation between saliva and serum SARS-CoV-2-specific IgG

Twenty-eight participants provided matched saliva and serum samples that were collected during the same visit (n=6 negative and n=22 RT-PCR confirmed SARS-CoV-2 infection matched saliva and serum samples). Antigen-specific IgG levels in matched saliva and serum samples were significantly correlated for all SARS-CoV-2 and SARS-CoV-1 antigens (**Figure 1**). Antigen-specific IgA in matched saliva and serum samples were modestly correlated with significance detected only for a subset of antigens: GenScript N, Sino Biol. N, Sino Biol. ECD, GenScript S1, and NAC SARS 2002 N (**Figure 2**). Antigen-specific IgM in matched saliva and serum samples were also significantly correlated for all SARS-CoV-2 and SARS-CoV-1 antigens, although the correlation was weaker than for IgG (**Figure 3**).

**Figure 1.**
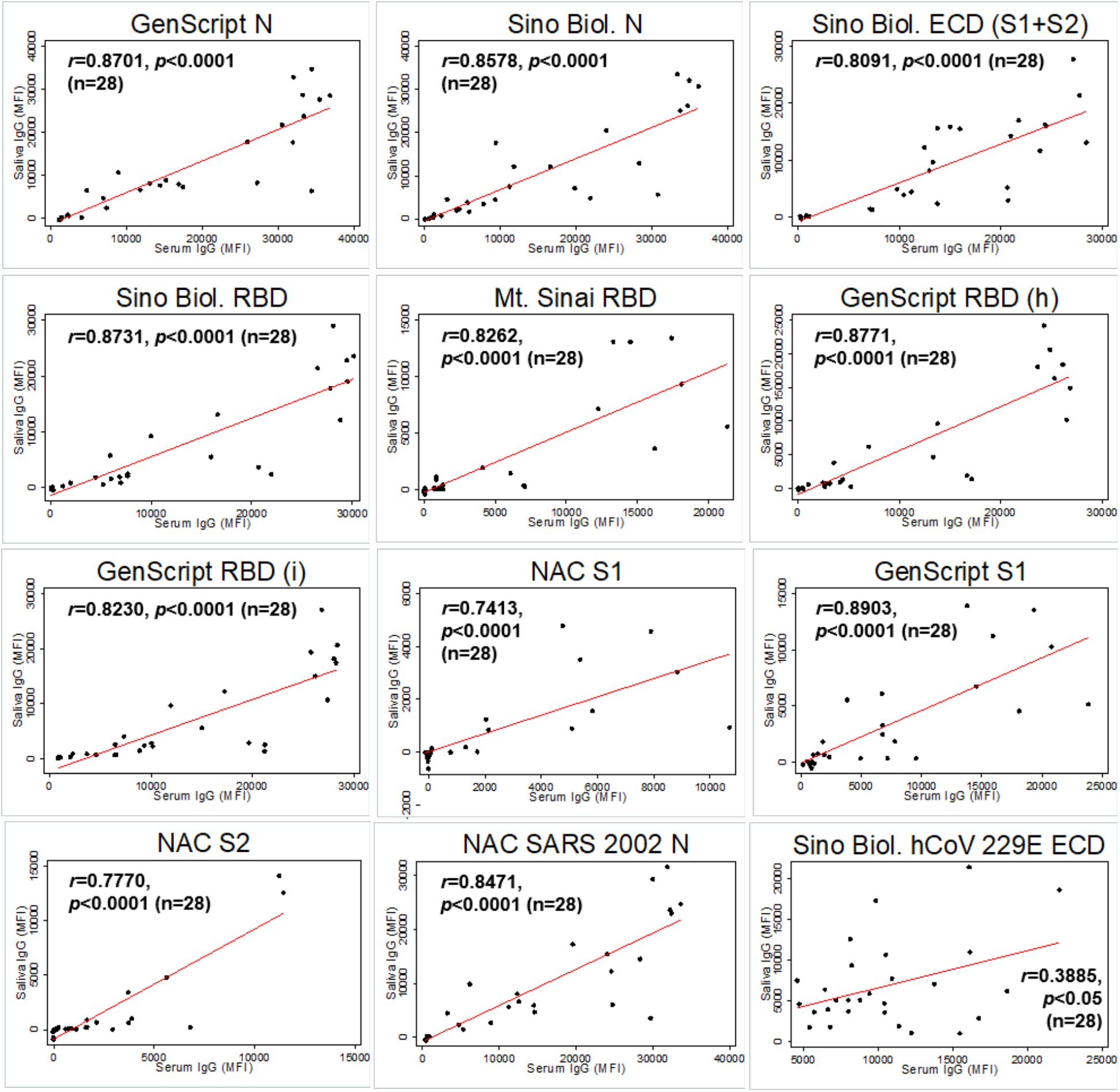
Correlation between saliva and serum SARS-CoV-2 antigen-specific IgG among matched saliva and serum samples (n=28). Pearson correlation coefficient is provided for each antigen-specific IgG. *p* values are provided for statistically significant correlations only (p<0.05). *Note*. Sino Biol.: Sino Biological; NAC: Native Antigen Company; N: nucleocapsid protein; ECD: S1: S1 subunit of spike protein; S2: S2 subunit of spike protein; ectodomain (S1 subunit+S2 subunit of the spike protein); RBD: receptor binding domain; (h): produced in human cell; (i): produced in insect cell; MFI=mean fluorescence intensity.

**Figure 2.**
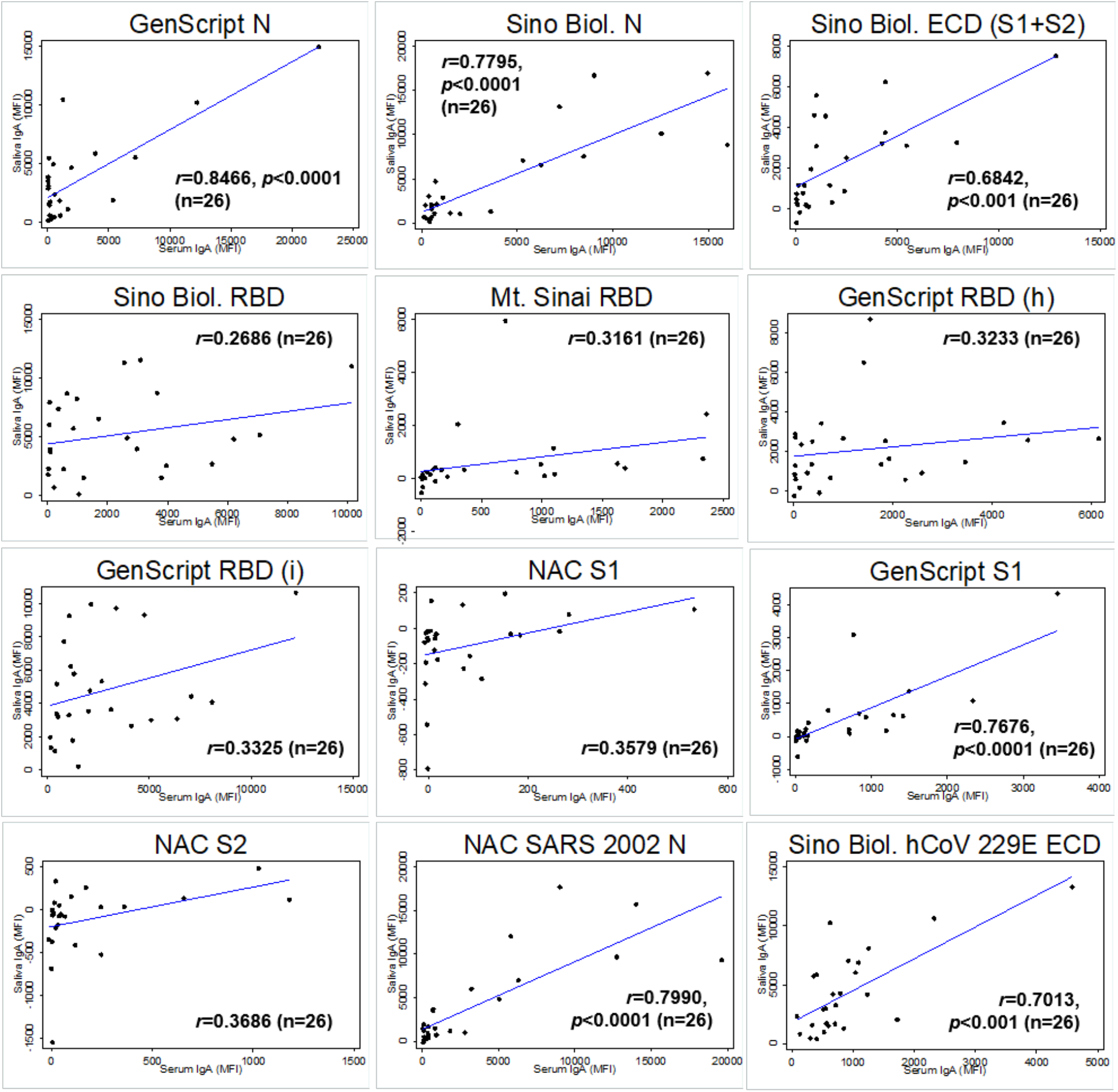
Correlation between saliva and serum SARS-CoV-2 antigen-specific IgA among matched saliva and serum samples (n=26). Pearson correlation coefficient is provided for each antigen-specific IgA. *p* values are provided for statistically significant correlations only (p<0.05). *Note*. Sino Biol.: Sino Biological; NAC: Native Antigen Company; N: nucleocapsid protein; ECD: S1: S1 subunit of spike protein; S2: S2 subunit of spike protein; ectodomain (S1 subunit+S2 subunit of the spike protein); RBD: receptor binding domain; (h): produced in human cell; (i): produced in insect cell; MFI=mean fluorescence intensity.

**Figure 3.**
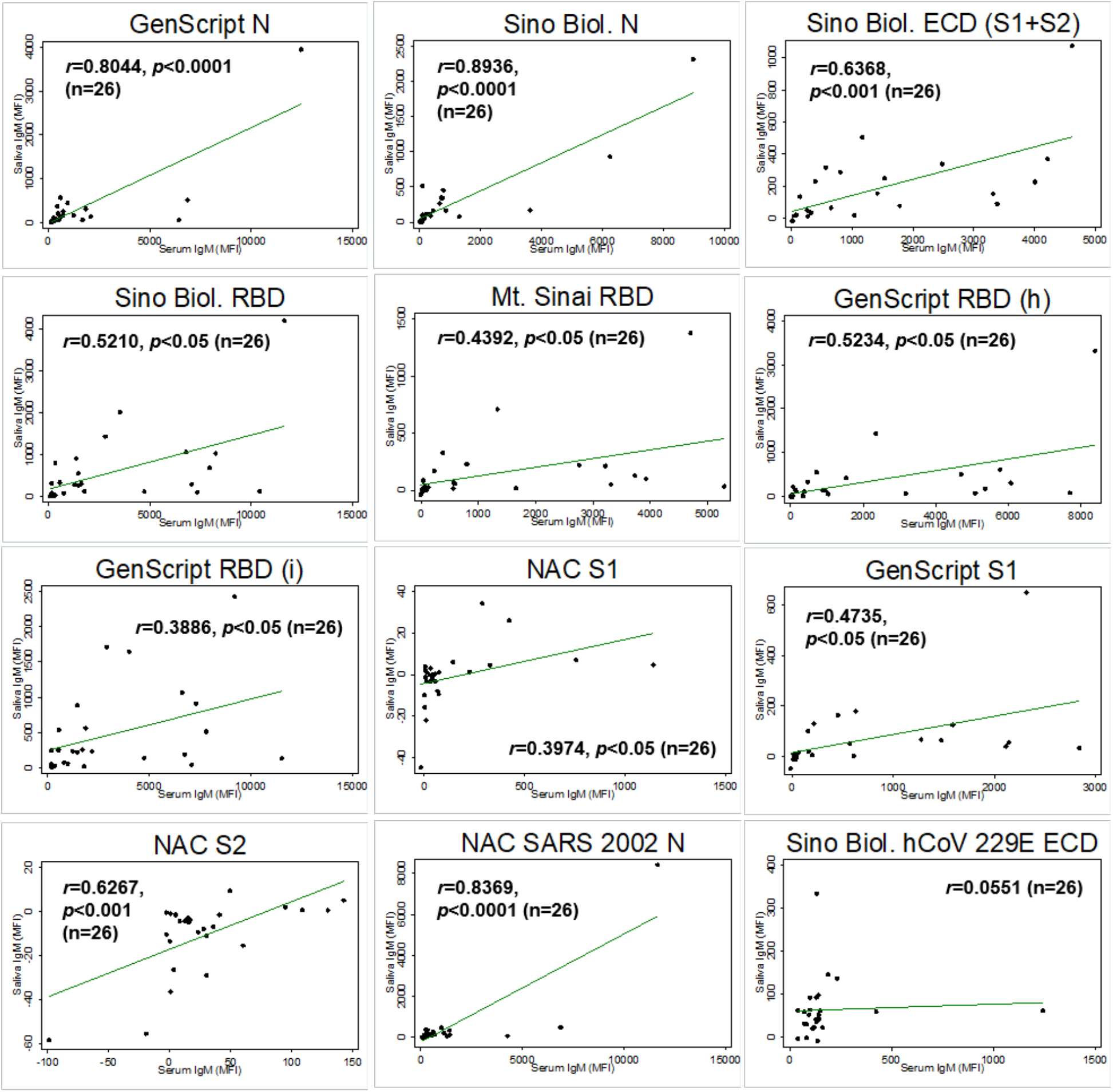
Correlation between saliva and serum SARS-CoV-2 antigen-specific IgM among matched saliva and serum samples (n=26). Pearson correlation coefficient is provided for each antigen-specific IgM. *p* values are provided for statistically significant correlations only (p<0.05). *Note*. Sino Biol.: Sino Biological; NAC: Native Antigen Company; N: nucleocapsid protein; ECD: S1: S1 subunit of spike protein; S2: S2 subunit of spike protein; ectodomain (S1 subunit+S2 subunit of the spike protein); RBD: receptor binding domain; (h): produced in human cell; (i): produced in insect cell; MFI=mean fluorescence intensity.

### Saliva: Sensitivity and specificity

In saliva, the sensitivity to detect SARS-CoV-2 infection increased among saliva samples collected ≥10 days post symptom onset compared to those collected <10 days post symptom onset, for all isotypes (IgG, IgA, and IgM)(Figure 4). The highest sensitivity (100%) was achieved with GenScript N-coupled beads in saliva samples collected ≥10 days post symptom onset. All (28/28) individuals with RT-PCR confirmed prior SARS-CoV-2 infection had salivary anti-GenScript N IgG levels above the cut-off (**Figure 4**). Specificity to classify negative saliva samples correctly ranged from 98% to 100% for SARS-CoV-2 IgG. Mt. Sinai’s RBD resulted in the highest specificity (100%). All (134/134) negative saliva samples resulted in MFI values below the cut-off (mean + 3 SD) for anti-Mt. Sinai RBD IgG levels. The highest combined sensitivity and specificity was achieved with GenScript N (100% sensitivity and 99% specificity at ≥10 days post symptom onset).

**Figure 4.**
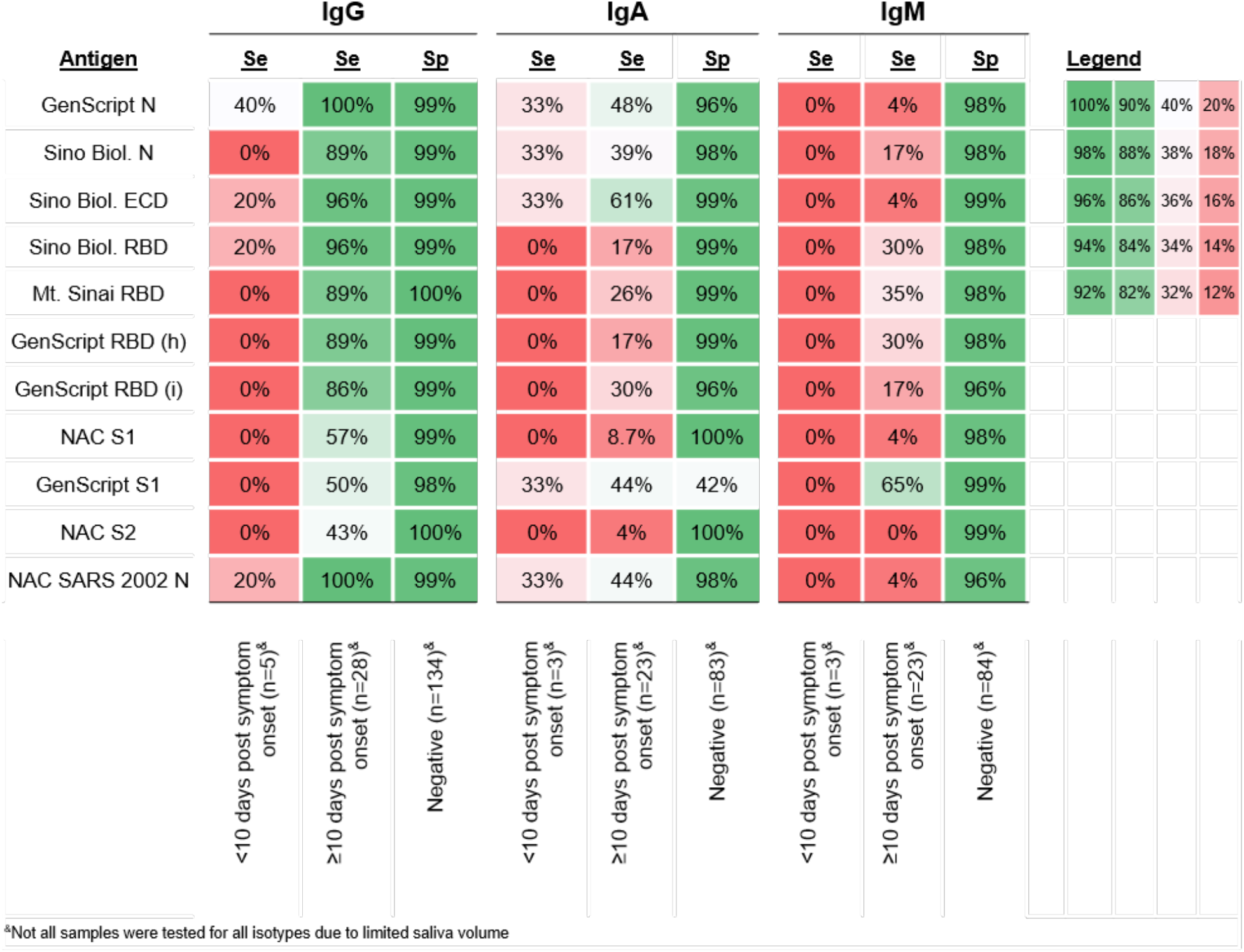
The sensitivity and specificity of each SARS-CoV-2 antigen-specific IgG, IgA, and IgM in saliva. Samples collected from individuals with RT-PCR confirmed prior SARS-CoV-2 infection are stratified into samples collected <10 days post symptom onset and samples collected ≥10 days post symptom onset. The average MFI of negative samples + 3 standard deviations was used to set the MFI cut off for each SARS-CoV-2 antigen-specific IgG, IgA, and IgM. Darker shades of green indicate higher whereas darker shades of red indicate lower sensitivity and specificity. *Note*. Sino Biol.: Sino Biological; NAC: Native Antigen Company; N: nucleocapsid protein; ECD: S1: S1 subunit of spike protein; S2: S2 subunit of spike protein; ectodomain (S1 subunit+S2 subunit of the spike protein); RBD: receptor binding domain; (h):produced in human cell; (i): produced in insect cell; Se: Sensitivity; Sp: specificity; MFI=mean fluorescence intensity.

While IgA and IgM against SARS-CoV-2 also remained equivalent or increased among saliva samples collected ≥10 days compared to <10 days post symptom onset, the sensitivity to detect prior SARS-CoV-2 infection remained low (Figure 4). For SARS-CoV-2 specific IgA, sensitivity ranged from 4% with NAC S2 to 61% with Sino Biol. ECD. For IgM, sensitivity ranged from 0% with NAC S2 to 65% with GenScript S1. Specificity for IgA ranged from 42% with GenScript S1 to 100% with NAC S1 and S2. The highest combined sensitivity and specificity for IgA was obtained with Sino Biol. ECD (61% sensitivity ≥10 days post symptom onset and 96% specificity). For IgM, specificity ranged from 96% (GenScript RBD [i]) to 99% (Sino Biol. ECD, GenScript S1, and NAC S2). The highest combined sensitivity and specificity for IgM was reached with GenScript S1 (65% sensitivity, 99% specificity).

### Serum: Sensitivity and specificity

In serum, the sensitivity to detect SARS-CoV-2 infection improved among serum samples collected ≥10 days compared to <10 days post symptom onset, for all isotypes (IgG, IgA, and IgM)(**Figure 5**). For anti-SARS-CoV-2 IgG, the highest sensitivity (92%) achieved with Mt. Sinai and Sino Biol. RBD using sera collected ≥10 days post symptom onset (96/104 samples from individuals with RT-PCR confirmed prior SARS-CoV-2 infection had IgG levels against these antigens above the cut-offs) (**Figure 5**). Specificity ranged from 96%-99% for anti-SARS-CoV-2 IgG. The highest combined sensitivity and specificity was achieved with Mt. Sinai’s RBD (92% sensitivity and 99% specificity at ≥10 days post symptom onset)

**Figure 5.**
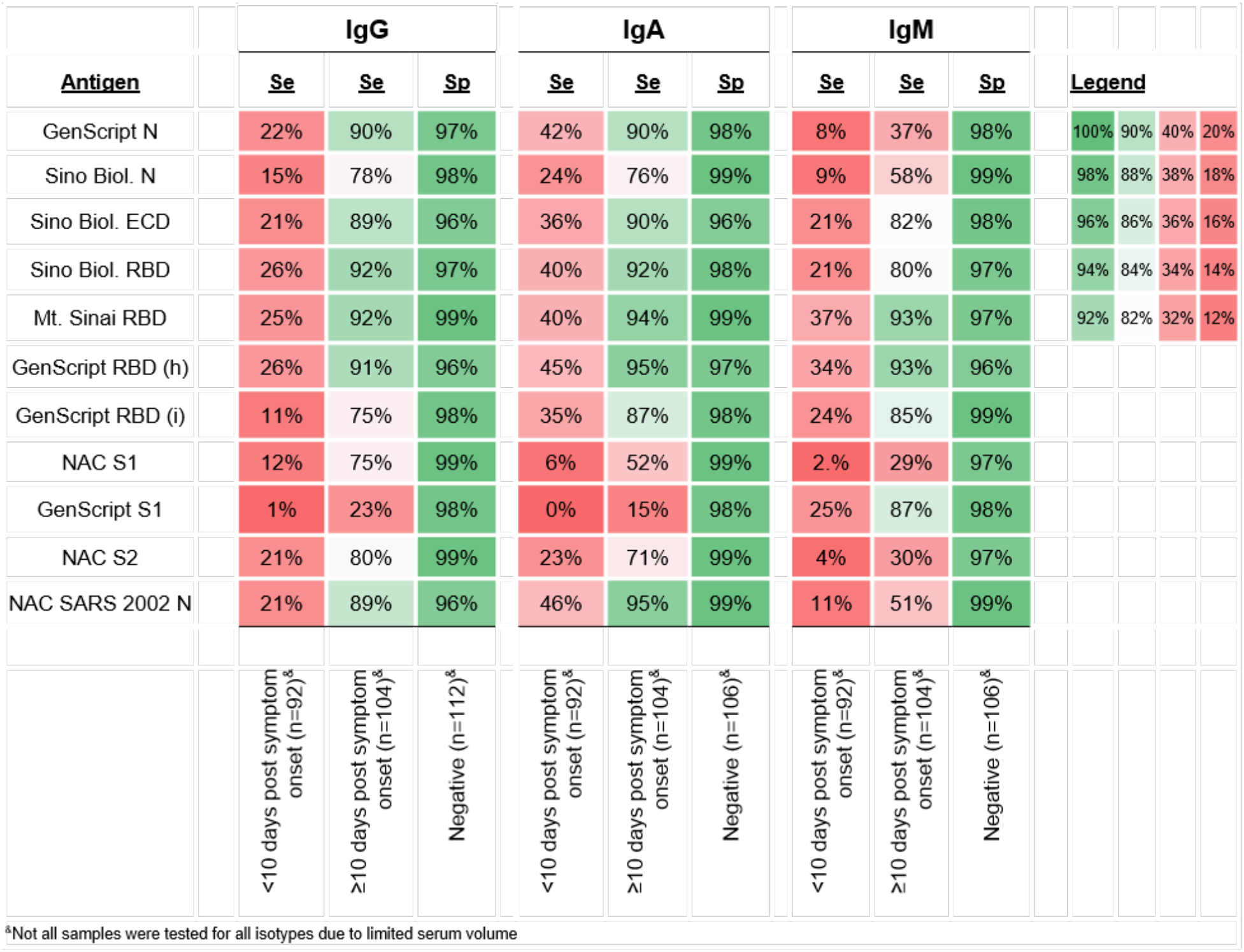
The sensitivity and specificity of each SARS-CoV-2 antigen-specific IgG, IgA, and IgM in serum. Samples collected from individuals with RT-PCR confirmed prior SARS-CoV-2 infection are stratified into samples collected <10 days post symptom onset and samples collected ≥10 days post symptom onset. The average MFI of negative samples + 3 standard deviations was used to set the MFI cut off for each SARS-CoV-2 antigen-specific IgG, IgA, and IgM. Darker shades of green indicate higher whereas darker shades of red indicate lower sensitivity and specificity. *Note*. Sino Biol.: Sino Biological; NAC: Native Antigen Company; N: nucleocapsid protein; ECD: S1: S1 subunit of spike protein; S2: S2 subunit of spike protein; ectodomain (S1 subunit+S2 subunit of the spike protein); RBD: receptor binding domain; (h):produced in human cell; (i): produced in insect cell; Se: Sensitivity; Sp: specificity; MFI=mean fluorescence intensity.

For anti-SARS-CoV-2 IgA and IgM, sensitivity ranged from 0% to 45% when using serum samples collected <10 days post-COVID-19 symptom onset; sensitivity was higher overall when detecting IgA compared to IgM. The sensitivity improved significantly with several antigens (predominantly RBD), when samples collected ≥10 days post-COVID-19 symptom onset were tested (**Figure 5**). When testing these sera, the highest sensitivity to detect IgA was reached using GenScript RBD (h) antigen (95%; 99/104 samples above the cutoff) but several additional antigens also performed with high sensitivity. In contrast, only two antigens (Mt. Sinai RBD and GenScript RBD [h]) in the assay reached sensitivities above 90% when detecting anti-SARS-CoV-2 IgM. Specificity ranged from 96%-99% for both anti-SARS-CoV-2 IgA and IgM. The highest combined sensitivity and specificity for detecting IgA and IgM was reached using Mt. Sinai’s RBD (as was the case for serum IgG) but also when using NAC’s SARS 2002 N antigen (**Figure 5**).

### Temporal kinetics of SARS-CoV-2 specific IgG, IgA, and IgM responses in serum compared to saliva

The temporal kinetics of antigen-specific IgG, IgA, and IgM responses in serum and in saliva are shown in Figure 6. Also shown are the cut-offs for each isotype (IgG, IgA, and IgM) in serum and in saliva (dashed lines). The temporal kinetics and magnitude of the antigen-specific IgG and IgA responses in saliva generally correlate with those detected in serum. The IgM response is significantly lower in magnitude (MFIs) in saliva compared to serum, which is expected and consistent with the lower relative concentration of total IgM in saliva compared to total IgA and IgG concentrations in saliva.

**Figure 6.**
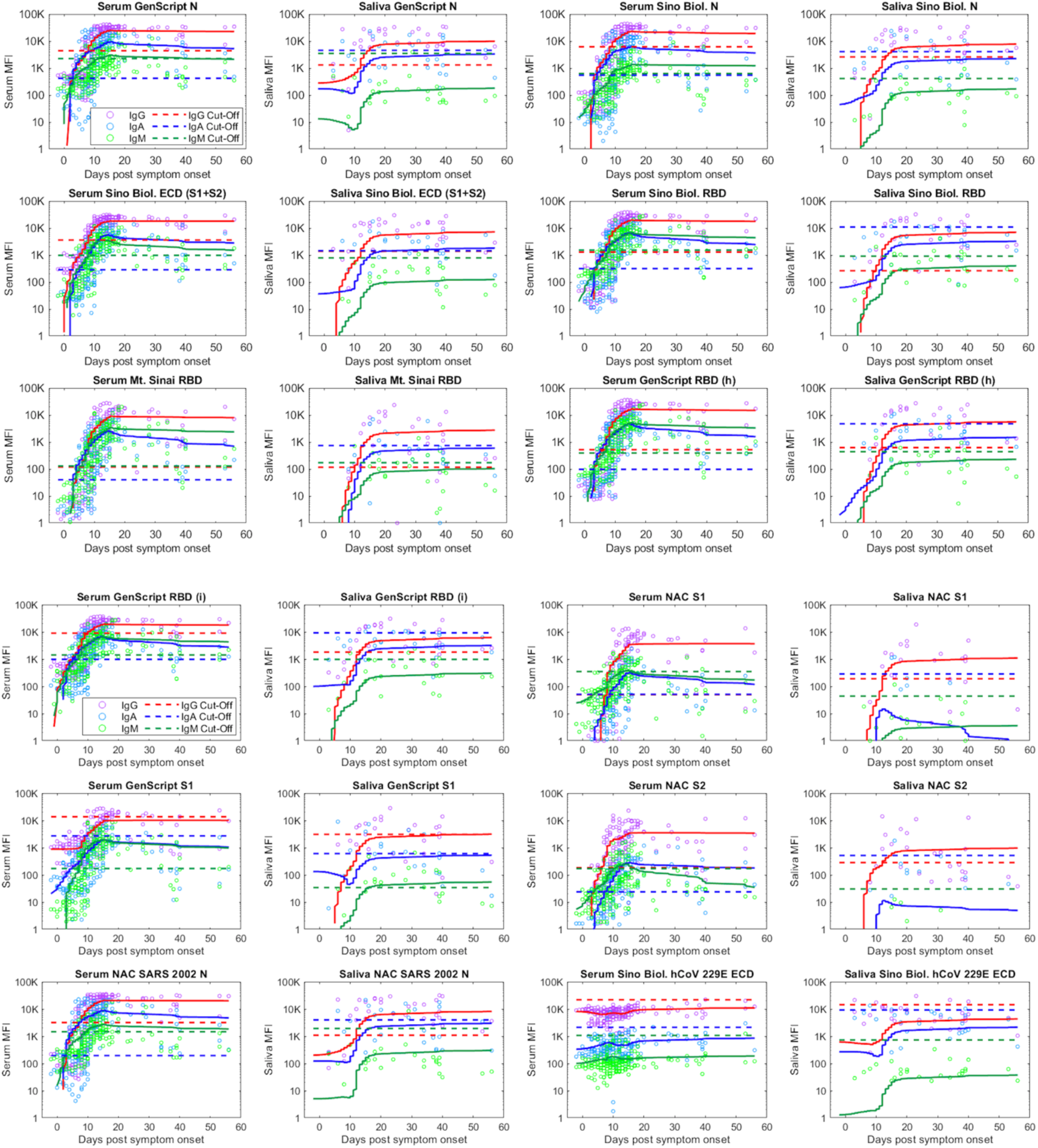
Comparison of saliva and serum SARS-CoV-2 antigen-specific IgG (red), IgA (blue), and IgM (green) responses vs. days post-COVID-19 symptom onset. The trajectories of IgG (red), IgA (blue), and IgM (green) responses are estimated using a LOESS curve. Dashed lines indicate cut off values for IgG (red), IgA (blue), and IgM (green). *Note*. Sino Biol.: Sino Biological; NAC: Native Antigen Company; N: nucleocapsid prot ein; ECD: S1: S1 subunit of spike protein; S2: S2 subunit of spike protein; ectodomain (S1 subunit+S2 subunit of the spike protein); RBD: receptor binding domain; (h): produced in human cell; (i): produced in insect cell; MFI=mean fluorescence intensity.

In serum, the SARS-CoV-specific IgA levels across individuals consistently cross the cut-off (dashed lines), thus indicating seroconversion, several days before IgG and IgM. IgG and IgM seroconversion in serum seem to occur approximately at the same time.

Even though saliva IgA levels increase closely after IgG levels, the SARS-CoV-2-specific IgA response often does not cross the cut-off, indicative of the low observed sensitivity. IgM levels in saliva are low and LOESS regression lines generally remain under the cut-off for most antigens in the multiplex assay. However, in saliva, the antigen-specific IgG response consistently crosses the cut-off around 10 days post symptom onset, i.e. approximately 15 days post infection, similar, to the time of IgG seroconversion in serum. The anti-SARS-CoV-2 IgG response in saliva thus appears to mimic seroconversion in serum.

### Reactivity of antibodies with SARS-CoV-1 and hCoV proteins following SARS-CoV-2 infection

We sought to evaluate reactivity of SARS-CoV-1 and hCoV proteins in samples from COVID-19 cases. For IgG, all convalescent phase saliva from COVID-19 cases (28/28; 100%) reacted with the NAC SARS 2002 N protein. Similarly, 89% and 95% of convalescent sera from COVID-19 cases reacted with the NAC SARS 2002 N protein for IgG and IgA, respectively. The median MFI for salivary IgG and IgA, and serum IgG, IgA, and IgM, to NAC SARS 2002 N was significantly elevated among samples collected ≥10 days post symptom onset compared to negatives (**Supplementary Table 1** and **Supplementary Table 2**). The median MFI for saliva and serum IgG and IgA to Sino Biol. hCoV 229E ECD was also elevated among samples collected ≥10 days post symptom onset compared to negatives (**Supplementary Table 1** and **Supplementary Table 2**). These results suggest that SARS-CoV-2 elicits cross-reactive antibodies to the closely related SARS-CoV-1, and that reactivity to Sino Biol. hCoV 229E ECD is very common in our study population, likely due to frequent human exposure to hCoVs.

### Intra- and inter-assay variability

Among 47 saliva samples assayed in duplicate on the same 96-well plate, the average intra-assay variability ranged from 3%-18% (CV%) (**Supplementary Table 3**). Among 47 saliva samples tested in duplicate on different 96-well plates on different days, the average inter-assay variability ranged from 5%-28% (CV%) (**Supplementary Table 3**).

## Discussion

Our results demonstrate that salivary SARS-CoV-2-specific IgG detection reflects the binding profile observed in serum. Salivary SARS-CoV-2-specific IgG can be used to detect a prior SARS-CoV-2 infection with high sensitivity and specificity. When saliva was collected ≥10 days post symptom onset, the anti-SARS-CoV-2 IgG assay detects SARS-CoV-2 infection with 100% sensitivity and 99% specificity (GenScript N) and/or with 89% sensitivity and 100% specificity (Mt. Sinai RBD). In addition, we demonstrate that the temporal kinetics of SARS-CoV-2-specific IgG responses in saliva are consistent with those observed in serum and indicate that most individuals seroconvert approximately 10 days after COVID-19 symptom onset or approximately two weeks post-presumed infection. Based on these results it is feasible to accurately measure the salivary IgG response to identify individuals with a prior SARS-CoV-2 infection. Our saliva-based multiplex immunoassay could serve as a non-invasive approach for accurate and large-scale SARS-CoV-2 “sero”-surveillance. Because saliva samples can be self-collected and mailed at ambient temperatures,^24^ a saliva antibody test could greatly increase the scale of testing—particularly among susceptible populations—compared to blood, and could clarify population immunity and susceptibility to SARS-CoV-2.

Matched saliva and serum samples demonstrate a significant correlation in SARS-CoV-2 antigen-specific IgG responses. An analysis of temporal kinetics of antibody responses in saliva following COVID-19 symptom onset revealed a congruence with those observed in serum, and a synchronous elevation of SARS-CoV-2 serum IgG and IgM responses, which has been reported in serum.^14,29-32^ In both saliva and serum, IgG rather than IgM was the first isotype to increase, mimicking a response consistent with the stimulation of existing, cross-reactive B cells, even though this is a novel coronavirus in these human populations. Both synchronous and classical antibody isotype responses have been previously reported following SARS-CoV-2 infection.^14,29-32^ Furthermore, IgG levels in saliva and serum tended to rise and cross the cut-off around day 10 post-COVID-19 symptoms onset, which is typically when individuals seek care from a healthcare provider for the first time. Therefore, salivary antibody testing could be used in combination with standard SARS-CoV-2 nucleic acid diagnostic testing to provide critical information about antibody positivity and temporal kinetics, which can be informative for patient trajectories and outcomes.

The sensitivity of our assay improved or remained the same among saliva and serum samples collected during convalescent phase (≥10 days post symptom onset) compared to acute phase (<10 days post symptom onset) for all SARS-CoV-2 antigen-specific IgG, IgA, and IgM. Saliva SARS-CoV-2 antigen-specific IgG peaked at 100% sensitivity, and serum SARS-CoV-2 antigen-specific IgG at 92% sensitivity (anti-Sino Biol. RBD IgG and anti-Mt. Sinai RBD IgG, respectively) among samples collected ≥10 days post SARS-CoV-2 symptom onset. Earlier studies have reported sensitivities for various SARS-CoV-2 IgG tests peaking at 82%-100% sensitivity among samples collected during convalescent phase of infection.^33-35^

While serum IgA and IgM peaked at 95% and 93% sensitivity, respectively, at >=10 days post symptom onset, saliva IgA and IgM reached a sensitivity of only 61% and 65%, respectively. The median MFI for most SARS-CoV-2 antigen-specific IgA and IgM responses in saliva were, however, significantly elevated at ≥10 days post symptom onset compared to negative control samples (Supplementary Table 1). One explanation for the low sensitivity observed in saliva for IgA and IgM may be due to the background signal-to-noise ratio for saliva SARS-CoV-2 antigen-specific IgA and IgM, which was greater than that observed for saliva IgG. Non-specific binding of salivary proteins, exogenous particles, non-specific antibodies, or cross-reactivity with other viruses could contribute to this background. Although we harvested GCF, which is enriched with blood transudate, because of size exclusion IgM antibodies are not abundant in saliva (12). Nevertheless, SARS-CoV-2 antigen-specific IgG responses in saliva performed with improved sensitivity and specificity compared to serum, peaking at 100% sensitivity ≥10 days post symptom onset for anti-GenScript N IgG and 100% specificity for anti-Mt. Sinai RBD IgG.

Virus infections often induce antibody responses that cross-react with related viruses, which can compromise the performance of serologic assays. Cross-reactivity may largely be attributable to the N protein and S2 subunit, which share 90% sequence homology with SARS-CoV-1.^31^ The RBD of the S protein is less conserved across beta-CoVs than the N protein and whole S protein, and many antibodies known to interact with SARS-CoV-1’s RBD do not interact with SARS-CoV-2’s RBD.^38^ For these reasons, we hypothesized that SARS-CoV-2 N would be highly sensitive and cross-react with antibodies following SARS-CoV-1 infection, whereas those against SARS-CoV-2 RBD would be more specific.^36^ We found that all (28/28; 100%) saliva samples from COVID-19 cases collected at ≥10 days post-symptom onset reacted with NAC SARS 2002 N in the IgG assay, indicating that SARS-CoV-2 infection can elicit cross-reactive IgG to closely related CoVs. Of course, this antigen could still be used for SARS-CoV-2 diagnostics, as the cross-reactivity would only be relevant if SARS-CoV-1 and SARS-CoV-2 were co-circulating in the same human population. We did not specifically evaluate whether common hCoVs elicit cross-reactive antibodies that could cause false positive results in our SARS-CoV-2 assay; however, we did include one hCoV antigen (hCoV-229E ECD) in the panel. Sera from early and late COVID-19 cases and negative control samples all reacted similarly to this antigen, which is consistent with a high prevalence of hCoV exposure in the general population.^39-41^ This also strongly suggests that our negative control sample population was highly exposed to hCoV and we would not have been able to achieve such clear discrimination between negative control and COVID-19 samples with other antigens in the multiplex panel if cross-reactivity was a significant issue.

This study has several limitations. First, our collection of saliva and serum samples was predominantly obtained from independent cohorts, and it contained 28 matched saliva and serum samples collected from the same participants at the same time. In future studies, the performance of this assay should be compared between saliva and serum in a large sample of matched saliva and serum samples. Second, all saliva data was cross sectional and we were not able to evaluate the temporal kinetics of saliva SARS-CoV-2 antibody responses using repeated measures within the same individual. Longitudinal analysis would allow us to evaluate the temporal kinetics and magnitude of SARS-CoV-2 IgG, IgA, and IgM responses, resolve synchronous vs. classical isotype responses (IgM followed by IgA followed by IgG) following SARS-CoV-2 infection.^42^

Additional investigation with convalescent phase saliva and sera are needed to determine the stability of SARS-CoV-2-specific IgG responses. Third, we did not have information on severity of SARS-CoV-2 disease from each participant in this study, and thus were not able to determine the impact of severity of infection on antibody responses.^29^ Prior studies suggest that antibody responses are slightly elevated among individuals with severe infection.^29,30,42^ Future analysis should determine how severity of infection, and infectious dose, modifies antibody responses. Fourth, we did not determine receiver operating characteristic (ROC)-optimized MFI cut offs in this analysis. However, the cut offs used in this study (average of negatives + three standard deviations) are conservative. Future analysis should identify ROC-optimized cut offs, which could improve the sensitivity and specificity of this saliva assay. Lastly, we did not have sociodemographic and medical history information for participants, and thus were not able to evaluate the relationship of age, sex, and other factors on antibody responses.

In future analysis, additional replicates should be used to assess intra- and inter-assay variability, and a lower limit of detection should be determined for each antigen. Furthermore, well characterized sera from other hCoV and zoonotic CoV infections should be used to address potential cross-reactivity of antibodies following SARS-CoV-1, MERS-CoV, hCoV-OC43, hCoV-HKU1, hCoV-229E, and hCoV-NL63 infection with SARS-CoV-2 proteins. Lastly, the performance of this saliva assay should be compared head-to-head with other clinically utilized antibody tests.

Saliva represents a practical, non-invasive alternative to NP, OP, blood, and stool-based diagnostic specimens for COVID-19 diagnostic testing. Recently, saliva collection via passive drool (instructing patients to spit into a sterile urine specimen collection cup) was shown to be more sensitive than NP specimens for SARS-CoV-2 RNA detection by RT-PCR in COVID-19 patients.^26^ Furthermore, the U.S. Food and Drug Administration recently granted emergency use authorization for a saliva-based nucleic acid test for SARS-CoV-2 that can be collected at home and mailed in for testing.^43^ Recognition of the advantages of saliva both for SARS-CoV-2 nucleic acid and antibody testing could accelerate goals for nationwide testing to surveil active and prior SARS-CoV-2 infections at the general population level.

This study demonstrates that SARS-CoV-2 antigen-specific antibody responses in saliva reflect those observed in serum, and that SARS-CoV-2 antigen-specific IgG can be used to accurately detect prior SARS-CoV-2 infection. We have developed and validated a saliva-based multiplex immunoassay and identified SARS-CoV-2 antigen-specific IgG responses that can detect prior SARS-CoV-2 infection with high sensitivity (anti-N IgG; 100% sensitivity, 99% specificity) and specificity (anti-RBD IgG; 89% sensitivity, 100% specificity) at ≥10 days post symptom onset. An accurate saliva-based antibody test for prior SARS-CoV-2 infection would greatly improve our ability to perform public health interventions in the current pandemic. This non-invasive method for comprehensive determination of prior SARS-CoV-2 infection will facilitate large-scale “sero”-surveillance to evaluate population immunity. As SARS-CoV-2 vaccine candidates progress through clinical trials, such non-invasive tests will be critical to identify immunity gaps and susceptible populations to inform targeted vaccination efforts, as well as companion diagnostics for vaccine trials.^44^ Furthermore, saliva assays can be used to monitor correlates of protection and the force of transmission in community-based settings, pre- and post-vaccination/prevention strategies, to determine the effectiveness of population-based interventions and direct future preventative strategies.

## Data Availability

Requests for de-identified data should be submitted to the authors.

## Acknowledgements

We would like to thank the participants who provided saliva and blood specimens that were used in this study to help develop and validate the multiplex SARS-CoV-2 antibody assays. We thank the faculty and staff team of The Hope Clinic for a dedicated effort in recruiting subjects and collecting specimens. We thank Dr. Yerun Zhu and Dr. Daniel Espinoza for assistance with laboratory operations

